# Asian Lung Cancer Absolute Risk Models for lung cancer mortality based on China Kadoorie Biobank

**DOI:** 10.1101/2022.04.22.22274185

**Authors:** Matthew T. Warkentin, Martin C. Tammemägi, Osvaldo Espin-Garcia, Sanjeev Budhathoki, Geoffrey Liu, Rayjean J. Hung

## Abstract

**Background:** Lung cancer is the leading cause of cancer mortality globally. Early detection through screening can markedly improve prognosis and prediction models can identify high-risk individuals for risk-based screening. However, most models have been developed in North American cohorts of smokers and much less is known about risk factors for never-smokers, which represent a growing proportion of lung cancers, particularly for Asian populations.

**Methods:** Based on the China Kadoorie Biobank, a population-based prospective cohort study of 512,639 adults age 30-79 recruited between 2004-2008 with up to 12 years of follow-up, we built an Asian Lung Cancer Absolute Risk Model (ALARM) for lung cancer mortality using flexible parametric survival models, separately for ever- and never-smokers, accounting for competing risks of all-other-cause mortality. Model performance was evaluated in a 25% hold-out test set using the time-dependent area under the receiver operating characteristic curve (AUC) and by comparing the model-predicted and observed risks for model calibration.

**Results:** Predictors assessed in the never-smoker lung cancer mortality model were age, sex, household income, lung function, history of emphysema/bronchitis, family history of cancer, personal cancer history, BMI, passive smoking, and indoor air pollution. The ever-smoker model additionally assessed smoking status (former vs. current), duration, and intensity. The 5-year AUC based on the hold-out test set for the never and ever-smoker models were 0.77 (95% CI: 0.73-0.80) and 0.81 (95% CI: 0.79-0.84), respectively. The maximum 5-year risk for never and ever smokers were 2.6% and 12.7%, respectively.

**Conclusions:** This study is among the first to develop and test risk models specifically for Asian populations, separately for never (ALARM-NS) and ever-smokers (ALARM-ES). Our models identify Asian never- and ever-smokers at high-risk of death due to lung cancer with a high degree of accuracy and may identify those with risks exceeding common eligibility thresholds who would likely benefit from lung cancer screening.

## Introduction

Lung cancer is the leading cause of cancer mortality globally. In 2020, there was an estimated 2.2 million incident lung cancers and 1.8 million deaths due to lung cancer, representing 11.4% and 18.0% of all cancer-related incidence and mortality, respectively (1). This corresponds to nearly 1 in 10 cancer diagnoses and 1 in 5 cancer deaths. The five-year survival proportion for lung cancer patients remains poor at only 10-20%, though this varies between countries (1). However, several large randomized trials in populations of predominantly European ancestry have demonstrated a significant reduction in lung cancer mortality when screening with low-dose helical computed tomography (2–5). Identifying high-risk individuals who are likely to benefit from lung cancer screening remains an important public health priority for reducing lung cancer mortality.

The United States Preventive Services Task Force (USPSTF) recommendations were recently updated, and suggest screening adults age 50 to 80 years, with 20 or more pack-year history of smoking, and who have quit smoking no more than 15 years prior (6). Using these criteria would fail to screen any light, long-term former, or never-smokers, which represent a growing proportion of all lung cancer diagnoses, particularly in Asian populations. The proportion of lung cancers among never-smokers varies geographically, with about 15% of lung cancers occurring among never-smokers in North America, and as much as 30-40% in Asian countries (7).

Absolute risk models have been shown to be superior in identifying high-risk individuals for lung cancer screening compared to the USPSTF criteria; however, these models have been primarily developed in North American cohorts of ever-smokers. The widely-used PLCO_m2012_ risk model (8) was developed in a cohort of smokers in the United States, which may not be generalizable to Asian populations due to potential differences in risk factors and baseline risk. Much less is known about the risk factors for never-smokers in Asian populations, within which never-smoker lung cancer is more common than for other racial groups. While approximately 54% of worldwide lung cancers occurred in Asian countries (1), currently there is no validated risk prediction model specifically for Asian populations. The goal of the current study was to develop and evaluate a lung cancer mortality risk prediction model, specifically for Asian populations, with separate models developed for ever- and never-smokers.

## Methods

### Study Participants

This study used data from the China Kadoorie Biobank (CKB) which has been described in detail previously (9). In brief, the CKB is a population-based prospective cohort of 512,639 adults age 30 to 79 recruited between 2004 and 2008 from 10 geographically defined regions in China with 5.1 million person-years of follow-up and detailed collections of epidemiologic data. We excluded any participants with a self-reported personal history of lung cancer within 5 years of baseline. Mortality status was collected through electronic linkage to regional mortality registries with up to 12 years of follow-up. Deaths due to lung cancer was the primary outcome in this study. Deaths from all other causes (hereafter referred to as “other cause mortality”), excluding deaths due to lung cancer, precludes lung cancer-attributable death and was considered as a competing risk of mortality. Central ethics approval for CKB was obtained from Oxford University and the China National Centres for Disease Control and local ethics approval was obtained for each recruitment site. All participants provided written informed consent. This specific project was approved by the Research Ethics Board at Mount Sinai Hospital.

### Statistical Analysis

#### Absolute risk models

In the competing risks settings, the probability of death from lung cancer occurring within a defined time period is estimated as a function of all relevant cause-specific hazards (10,11). To estimate the probability of lung cancer mortality in the presence of the competing risk of other-cause mortality, we separately modeled the cause-specific hazards for death from lung cancer and death from other causes (i.e., deaths attributed to causes other than lung cancer). We estimated the five-year probability of lung cancer mortality using the cumulative incidence function which accounts for both hazard functions, conditional on a set of risk factors (See Supplementary Methods for more details).

We modeled the cause-specific hazards using flexible parametric survival models (i.e., Royston-Parmar models (12)) on the cumulative hazard scale to estimate baseline hazards and cause-specific hazard ratios for predictor effects (e.g., clinico-epidemiological and spirometry data). Models were fit separately for never and ever-smokers using complete-case analysis. We fit models on the time-since-entry timescale using restricted (natural) cubic splines with internal knots placed at the quantiles of the log uncensored cause-specific event-time distributions. Models were fit using the flexsurv package in R (13).

Asian Lung Cancer Absolute Risk Models (ALARM) were built separately for never (ALARM-NS) and ever-smokers (ALARM-ES). We used stratified random sampling to split the data into training and testing sets, maintaining similar proportions of the outcome, separately for never- and ever-smokers. Models were fitted in the training (75%) data, and the hold-out testing (25%) data were used to estimate out-of-sample performance for internal model validation.

For the lung cancer mortality models, potential predictors were selected based on their known associations with lung cancer or by improving model performance. ALARM-NS included age, sex, body mass index, family history of cancer, personal cancer history (excluding lung cancers within 5 years of enrolment), lung function (forced expiratory volume in 1 second (FEV1) / forced vital capacity (FVC)), personal history of emphysema or bronchitis, and household income (6 levels). ALARM-ES included these variables with the addition of smoking status (current vs. former), smoking duration (years), and smoking intensity (cigarettes / day). Cooking fuel exposure was computed as a composite of cooking fuel type (gas or electricity vs. all others) and duration of usage (years of usage at current and previous residences). Numeric variables were evaluated for potential non-linear relationships and time-dependent effects. Age was included in the final models as the logarithm of age, and BMI was included as quadratic (BMI and BMI^2^). The other cause mortality model included age, sex, and smoking status (never, former, current) as covariates. We report the cause-specific hazard ratios (HR) and 95% confidence intervals (CI). All statistical analyses were done using R version 4.0.5 (14).

#### Model performance

The performance of the prediction models was assessed in two complementary ways: (1) how closely model-predicted risks corresponded to observed risks (calibration) and, (2) the ability of the model to assign higher predicted risks to those who died of lung cancer, or died of lung cancer at an earlier time (discrimination).

Model calibration was assessed by comparing the observed five-year risks to model-predicted (expected) five-year risks, separately for never-smokers and ever-smokers. Graphical assessment was performed by comparing risks within binned risk groups. The observed five-year risks were estimated based on the non-parametric Aalen-Johansen estimator for the cumulative incidence function (15). The model-predicted five-year risks were estimated based on the absolute risk models described above. Ideal calibration corresponds to points falling along the diagonal line in a calibration plot with a slope of 1 (i.e., the identity line). The ratio of expected to observed deaths (E/O) and difference in expected and observed deaths (E-O) per 100,000 are reported for the never and ever-smoker models.

Discrimination was measured by the area under the time-dependent receiver operating characteristic curve (AUC). We used the definition of the time-dependent ROC for competing risks described in Blanche *et al* (17). We constructed 95% confidence intervals for the time-dependent AUC and calibration metrics using a percentile-based approach with 1000 bootstrap resamples. Calibration and discrimination metrics are reported for the hold-out test data only.

## Results

Basic demographic characteristics for the CKB cohort according to vital status and by smoking status are presented in **Table 1** and **Supplemental Table 1**, respectively, based on the total length of follow-up. In general, those who died from lung cancer were older at baseline (62.2 vs. 52.0 years), were more likely to be male (64% vs. 41%), and were more likely to be current or former smokers with extensive smoking history, when compared with those still alive.

**Table 1.** Distribution of demographic characteristics for China Kadoorie Biobank based on mortality status. Means and standard deviations are reported for numeric variables, and frequencies and proportions for categorical variables.

|  | Alive<br>No. (%) | Lung cancer mortality<br>No. (%) | All-cause mortality <sup>a</sup><br>No. (%) | Total<br>No. (%) |
| --- | --- | --- | --- | --- |
| N | 480,115 | 2,754 | 29,770 | 512,639 |
| Age (years) |  |  |  |  |
| mean [sd] | 51.3 [10.4] | 62.2 [9.1] | 62.4 [9.8] | 52.0 [10.7] |
| Length of follow-up (years) |  |  |  |  |
| mean [sd] | 10.3 [1.1] | 4.9 [2.3] | 4.8 [2.4] | 9.9 [1.8] |
| Sex |  |  |  |  |
| Female | 288,874 (60%) | 991 (36%) | 12,632 (42%) | 302,497 (59%) |
| Male | 191,241 (40%) | 1,763 (64%) | 17,138 (58%) | 210,142 (41%) |
| Family history of cancer |  |  |  |  |
| No | 397,851 (83%) | 2,253 (82%) | 25,228 (85%) | 425,332 (83%) |
| Yes | 82,264 (17%) | 501 (18%) | 4,542 (15%) | 87,307 (17%) |
| Personal cancer history |  |  |  |  |
| No | 478,187 (>99%) | 2,724 (99%) | 29,225 (98%) | 510,136 (>99%) |
| Yes | 1,928 (<1%) | 30 (1%) | 545 (2%) | 2,503 (<1%) |
| Emphysema or bronchitis |  |  |  |  |
| No | 469,102 (98%) | 2,553 (93%) | 27,696 (93%) | 499,351 (97%) |
| Yes | 11,013 (2%) | 201 (7%) | 2,074 (7%) | 13,288 (3%) |
| Smoking status |  |  |  |  |
| Never | 330,283 (69%) | 1,060 (38%) | 15,274 (51%) | 346,617 (68%) |
| Former | 32,901 (7%) | 415 (15%) | 4,652 (16%) | 37,968 (7%) |
| Current | 116,931 (24%) | 1,279 (46%) | 9,844 (33%) | 128,054 (25%) |
| Smoking Intensity (cigs/day) <sup>b</sup> |  |  |  |  |
| mean [sd] | 17.9 [10.8] | 19.0 [11.2] | 16.7 [11.1] | 17.8 [10.8] |
| Smoking duration (years) <sup>b</sup> |  |  |  |  |
| mean [sd] | 22.7 [7.3] | 22.2 [7.4] | 23.3 [8.8] | 22.7 [7.5] |
| Time since quitting (years) <sup>c</sup> |  |  |  |  |
| mean [sd] | 8.7 [8.5] | 8.2 [8.5] | 8.5 [8.9] | 8.7 [8.5] |
| FEV1/FVC (%) |  |  |  |  |
| mean [sd] | 84.8 [8.2] | 80.6 [10.5] | 80.4 [11.4] | 84.5 [8.5] |
| Body mass index (kg/m<sup>2</sup>) |  |  |  |  |
| mean [sd] | 23.7 [3.3] | 22.8 [3.6] | 23.0 [3.8] | 23.7 [3.4] |
| Household income <sup>d</sup> |  |  |  |  |
| <2500 yuan | 30,566 (7%) | 236 (9%) | 3,822 (14%) | 34,624 (7%) |
| 2,500-4,999 yuan | 87,540 (19%) | 502 (19%) | 6,519 (24%) | 94,561 (19%) |
| 5,000-9,999 yuan | 139,550 (30%) | 829 (32%) | 8,553 (32%) | 148,932 (30%) |
| 10,000-19,999 yuan | 121,086 (26%) | 647 (25%) | 4,948 (18%) | 126,681 (25%) |
| 20,000-34,999 yuan | 88,968 (19%) | 404 (15%) | 2,932 (11%) | 92,304 (19%) |
| ≥35,000 yuan | 139,550 (30%) | 829 (32%) | 8,553 (32%) | 148,932 (30%) |
| Cooking fuel exposure <sup>e</sup> |  |  |  |  |
| None | 191,331 (40%) | 1,242 (45%) | 11,803 (40%) | 204,376 (40%) |
| Low | 76,064 (16%) | 443 (16%) | 4,321 (15%) | 80,828 (16%) |
| Medium | 143,175 (30%) | 602 (22%) | 6,474 (22%) | 150,251 (29%) |
| High | 69,545 (14%) | 467 (17%) | 7,172 (24%) | 77,184 (15%) |

Abbreviations: FEV1, forced-expiratory volume, 1-second; FVC, forced vital capacity; sd, standard deviation.
<sup>a</sup> All-cause mortality included all causes of death except death attributable to lung cancer.
<sup>b</sup> Average smoking intensity and age at smoking initiation includes current and former smokers; never smokers are excluded.
<sup>c</sup> Years since smoking cessation only includes former smokers; current and never smokers are excluded.
<sup>d</sup> Approximate equivalents in USD (\$), rounded to the nearest dollar: <384, 384-767, 767-1,537, 1,537-3,075, 3,075-5,381, and ≥5381.
<sup>e</sup> Cooking fuel exposure groups were formed based on 25<sup>th</sup> and 75<sup>th</sup> percentiles of the cumulative cooking exposure distribution. Cumulative cooking exposure was calculated based on the self-reported frequency of exposure to potentially harmful cooking fuels (i.e., coal, wood, or other, as compared to gas or electricity).

In total, three separate flexible parametric cause-specific hazard models were fitted for: (1) lung cancer mortality for never-smokers, (2) lung cancer mortality for ever-smokers, and (3) all-other-cause mortality. The two smoking status-specific absolute risk models are formed based on models 1 and 3 (ALARM-NS) and models 2 and 3 (ALARM-ES). The cause-specific hazard ratios and 95% confidence intervals for the lung cancer mortality models are reported in **Table 2** and for all-other-cause mortality in **Supplemental Table 2**.

**Table 2.**
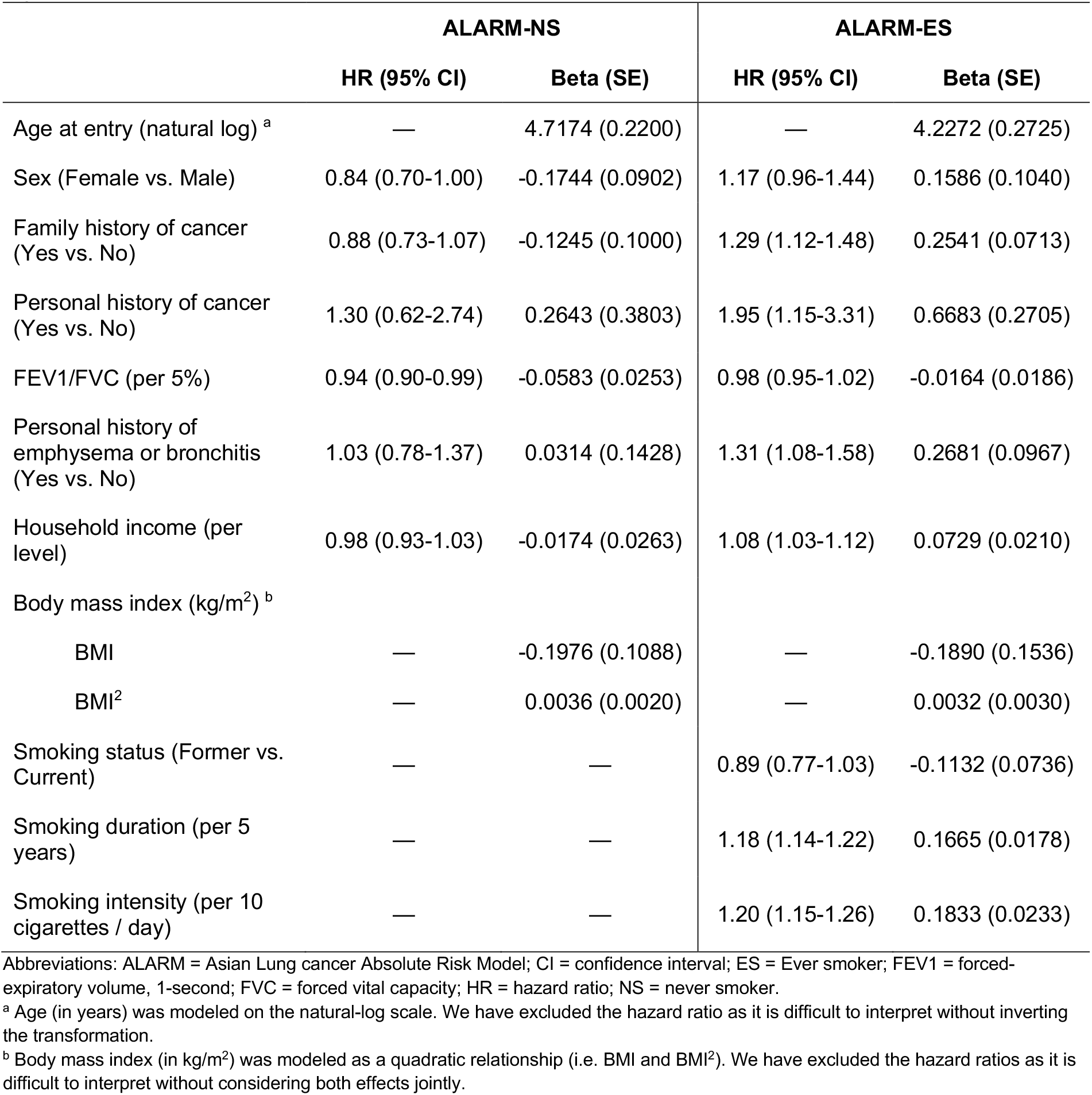
Estimates from lung cancer mortality flexible parametric survival models fit separately for never-smokers (ALARM-NS) and ever-smokers (ALARM-ES). Lung cancer cause-specific hazard ratios with 95% confidence intervals and beta coefficients (log hazard ratios) with standard errors are reported.

|  | ALARM-NS |  | ALARM-ES |  |
| --- | --- | --- | --- | --- |
|  | HR (95% CI) | Beta (SE) | HR (95% CI) | Beta (SE) |
| Age at entry (natural log) <sup>a</sup> | — | 4.7174 (0.2200) | — | 4.2272 (0.2725) |
| Sex (Female vs. Male) | 0.84 (0.70-1.00) | -0.1744 (0.0902) | 1.17 (0.96-1.44) | 0.1586 (0.1040) |
| Family history of cancer (Yes vs. No) | 0.88 (0.73-1.07) | -0.1245 (0.1000) | 1.29 (1.12-1.48) | 0.2541 (0.0713) |
| Personal history of cancer (Yes vs. No) | 1.30 (0.62-2.74) | 0.2643 (0.3803) | 1.95 (1.15-3.31) | 0.6683 (0.2705) |
| FEV1/FVC (per 5%) | 0.94 (0.90-0.99) | -0.0583 (0.0253) | 0.98 (0.95-1.02) | -0.0164 (0.0186) |
| Personal history of emphysema or bronchitis (Yes vs. No) | 1.03 (0.78-1.37) | 0.0314 (0.1428) | 1.31 (1.08-1.58) | 0.2681 (0.0967) |
| Household income (per level) | 0.98 (0.93-1.03) | -0.0174 (0.0263) | 1.08 (1.03-1.12) | 0.0729 (0.0210) |
| Body mass index (kg/m<sup>2</sup>) <sup>b</sup> |  |  |  |  |
| BMI | — | -0.1976 (0.1088) | — | -0.1890 (0.1536) |
| BMI<sup>2</sup> | — | 0.0036 (0.0020) | — | 0.0032 (0.0030) |
| Smoking status (Former vs. Current) | — | — | 0.89 (0.77-1.03) | -0.1132 (0.0736) |
| Smoking duration (per 5 years) | — | — | 1.18 (1.14-1.22) | 0.1665 (0.0178) |
| Smoking intensity (per 10 cigarettes / day) | — | — | 1.20 (1.15-1.26) | 0.1833 (0.0233) |

Abbreviations: ALARM = Asian Lung cancer Absolute Risk Model; CI = confidence interval; ES = Ever smoker; FEV1 = forced-expiratory volume, 1-second; FVC = forced vital capacity; HR = hazard ratio; NS = never smoker.
<sup>a</sup> Age (in years) was modeled on the natural-log scale. We have excluded the hazard ratio as it is difficult to interpret without inverting the transformation.
<sup>b</sup> Body mass index (in kg/m<sup>2</sup>) was modeled as a quadratic relationship (i.e. BMI and BMI<sup>2</sup>). We have excluded the hazard ratios as it is difficult to interpret without considering both effects jointly.

Age (on the log scale) increased the risk of lung cancer mortality for both never and ever smokers. BMI was included in the model as quadratic terms to capture the potential non-linear relationship between BMI and lung cancer mortality (18,19). Female sex was found to be protective for lung-cancer mortality in never-smokers (HR: 0.84, 95% CI: 0.70-1.00), but a modest risk factor in ever-smokers (HR: 1.17, 95% CI: 0.96-1.44). Family history of any type of cancer was a risk factor for ever-smoker death due to lung cancer (HR: 1.29, 95% CI: 1.12-1.48), but not for never-smokers. Personal cancer history increased hazard of lung cancer mortality in never (HR: 1.30, 95% CI: 0.62-2.74) and ever smokers (HR: 1.95, 95% CI: 1.15-3.31). For every 5% increase in lung function performance (FEV1/FVC), the hazard of lung cancer mortality was reduced by 6% and 2%, for never and ever smokers, respectively. In addition, a self-reported history of emphysema or bronchitis had HR of 1.03 (95% CI: 0.78-1.37) and 1.31 (95% CI: 1.08-1.58) for never and ever-smokers, respectively. Cooking fuel exposure was not found to improve model performance and was therefore not included from the final models.

ALARM-ES further included several smoking variables. When compared to current smokers, former smokers had a lower hazard of lung cancer morality (HR: 0.89, 95% CI: 0.77-1.03). For every 5 additional years of smoking, the hazard of lung cancer mortality increased by 18% (HR: 1.18, 95% CI: 1.14-1.22). For every additional 10 cigarettes smoked per day (approximately equivalent to half a pack), the hazard of lung cancer mortality was increased by 20% (HR: 1.20, 95% CI: 1.15-1.26). Non-linear relationships and two-way interactions were explored for smoking variables but did not contribute to model improvement and were not included in the final models.

We estimated the five-year absolute risk of lung cancer mortality (i.e., cumulative incidence of death from lung cancer), conditional on a participant’s risk factor profile. The distribution of risks for never- and ever-smokers are presented in **Supplemental Figure 1**. The maximum five-year predicted risk of lung cancer mortality was 12.7% for ever-smokers and 2.6% for never-smokers. According to our models, 8.1% of ever-smokers and <1% of never-smokers in the CKB would be eligible for screening assuming a 1.5% five-year risk threshold.

Calibration plots comparing model-predicted and observed five-year risks across risk quantiles, separately for ALARM-NS and ALARM-ES, are presented in **Supplemental Figure 2**. Both models show very good calibration in the hold-out test data. Additional calibration metrics overall are presented in **Supplemental Table 3**. The five-year time-dependent AUC in the hold-out test set 0.77 (95% CI: 0.73-0.80) and was 0.81 (95% CI: 0.79-0.84) for ALARM-NS and ALARM-ES models, respectively. The time-dependent receiver operating characteristic curves for the 25% hold-out test data are presented in **Figure 1**.

**Figure 1.**
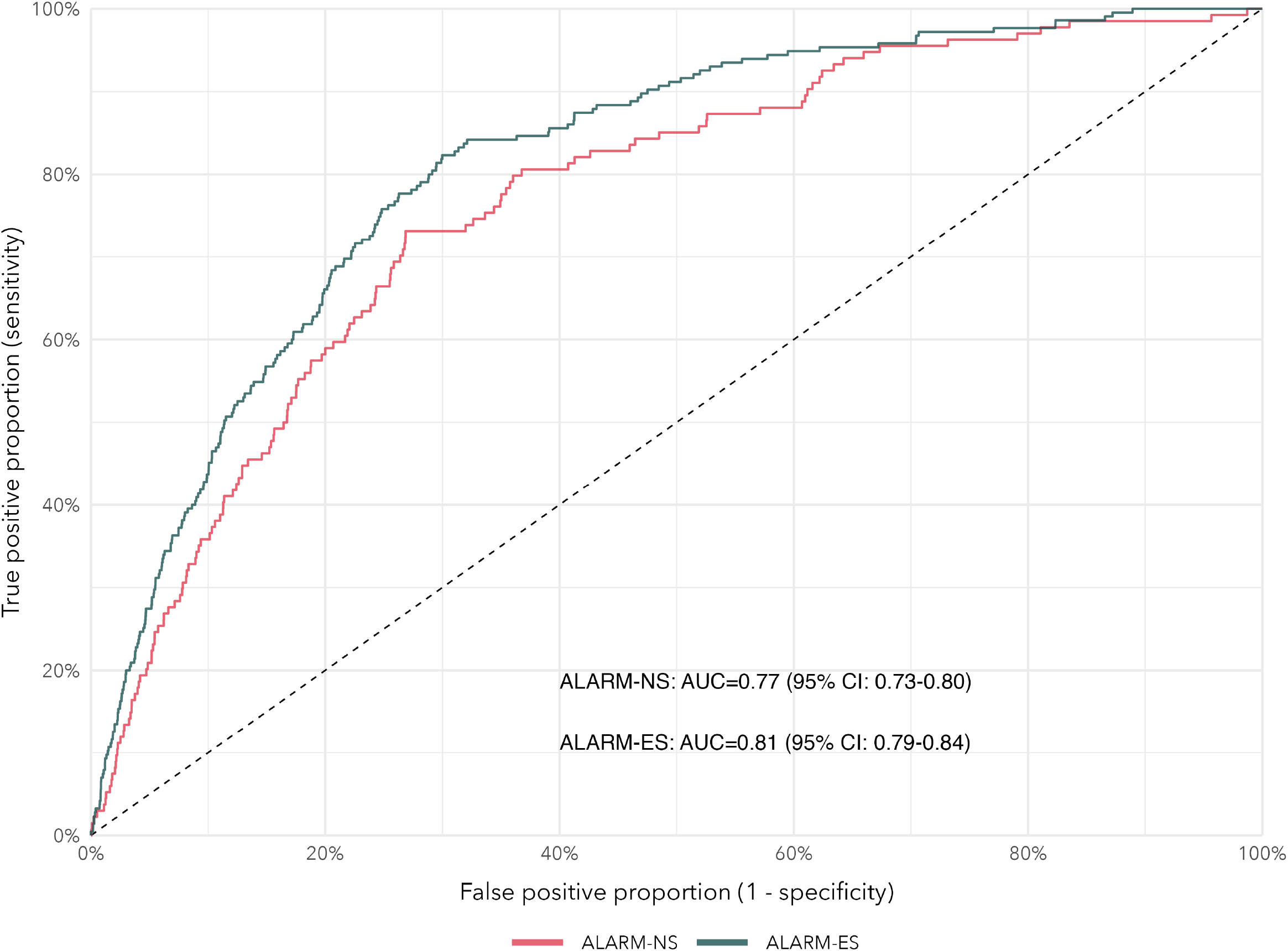
Five-year receiver operating characteristic (ROC) curves and area under the curves (AUC) for the 25% hold-out test data, separately for ALARM-NS (never-smokers) and ALARM-ES (ever-smokers).

Absolute risk trajectories for average and high-risk current and former smokers are presented in **Figure 2** according to lung function performance and smoking intensity. Lung cancer mortality risks are higher for those with suboptimal lung function and higher smoking intensity. Absolute risk trajectories for never-smokers are presented in **Figure 3**, separately for men and women according to lung function. Lung cancer mortality risk is higher for suboptimal lung function and for men. Contour plots for absolute risk according to smoking intensity, age at smoking initiation, and lung function for a theoretical average or high-risk current and former smokers are shown in **Supplemental Figure 3**. Contour plots for a theoretical average or high-risk never-smoker according to age, FEV1/FVC, and sex are shown in **Figure 4**, with risks as high as 2.25% to 2.50% for the highest risk profile.

**Figure 2.**
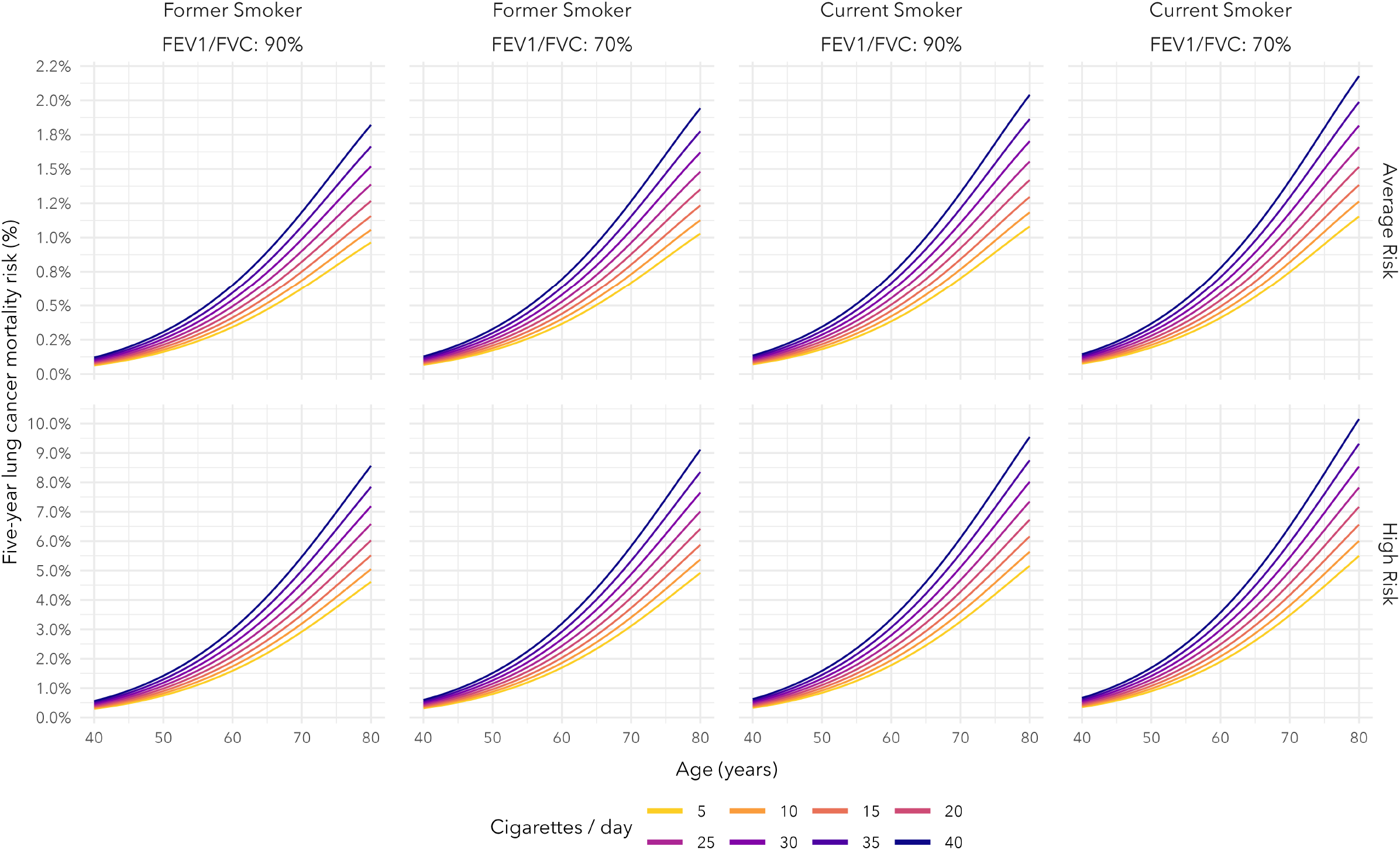
Five-year absolute risk trajectories for lung cancer mortality based on ALARM-ES for current and former smokers for varying smoking intensity (cigarettes per day) and lung function (FEV1/FVC), for average risk and high risk profiles. An average risk profile is defined as having the average covariate value for all predictors other than those varied, and a high risk profile is defined as having the highest risk covariate value observed in the CKB (based on the direction of effect) for all predictors other than those varied.

**Figure 3.**
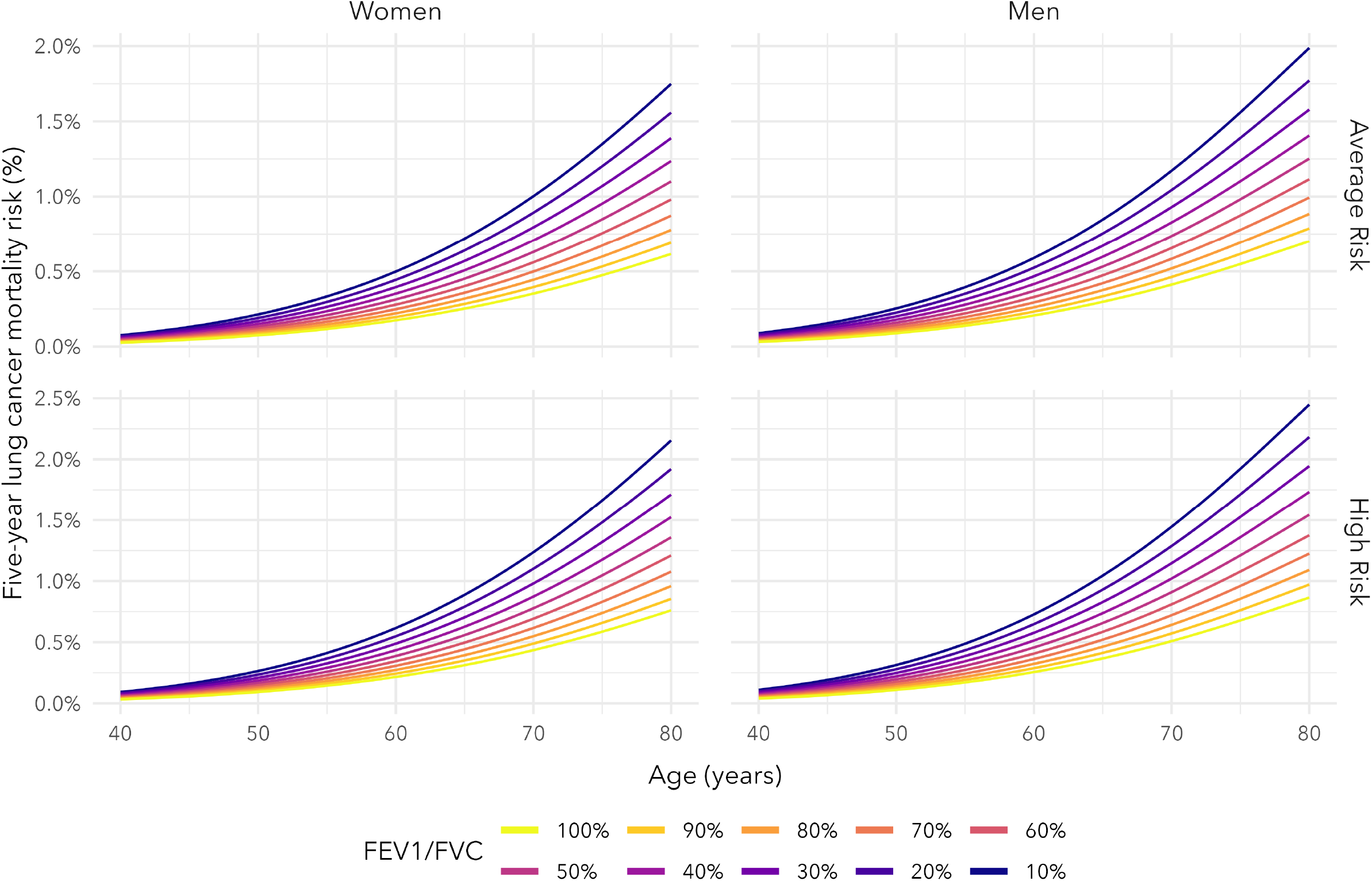
Five-year absolute risk trajectories for lung cancer mortality based on ALARM-NS for never smoker men and women across levels of FEV1/FVC, for average risk and high risk profiles. An average risk profile is defined as having the average covariate value for all predictors other than those varied, and a high risk profile is defined as having the highest risk covariate value observed in the CKB (based on direction of effect) for all predictors other than those varied.

**Figure 4.**
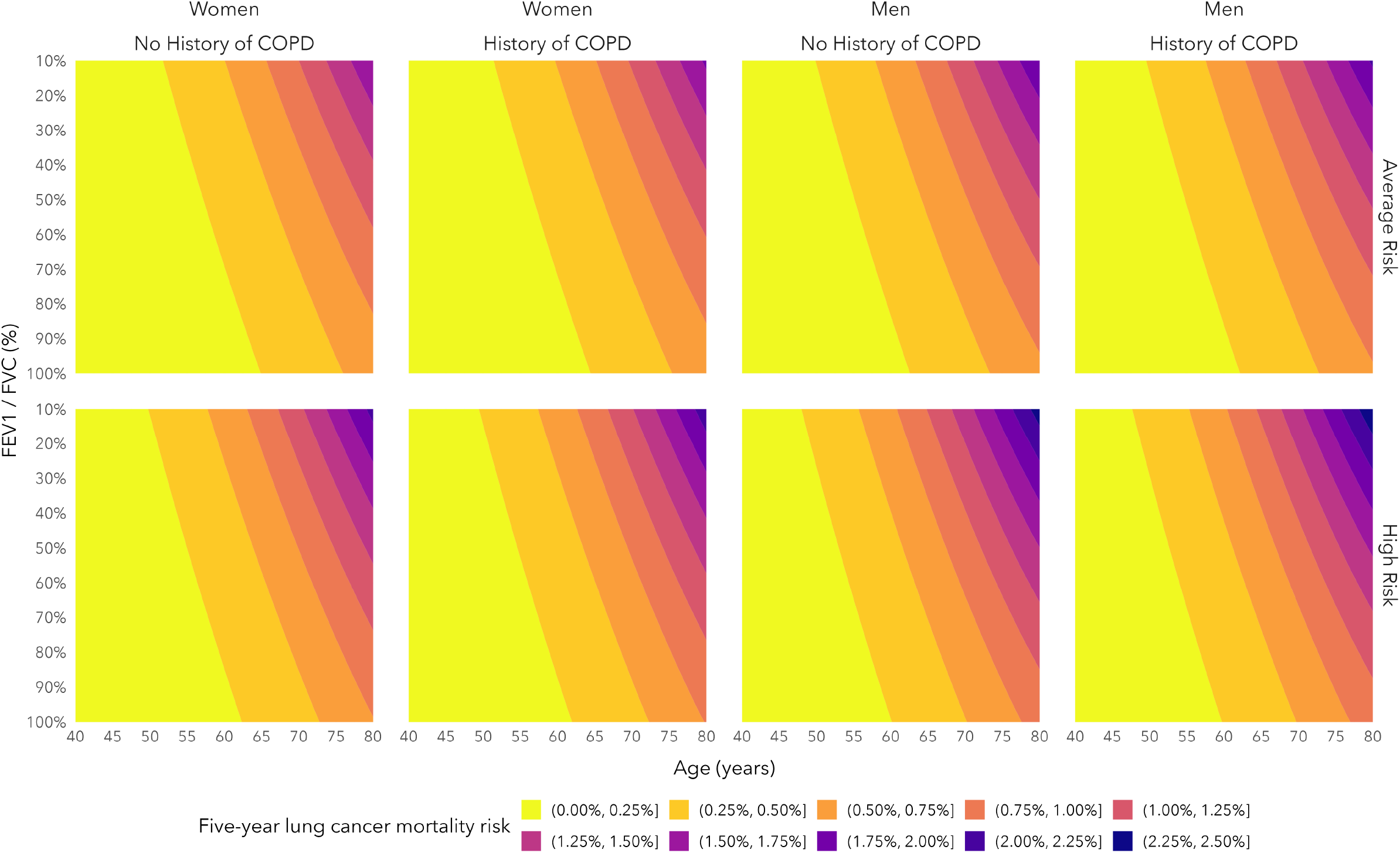
Five-year absolute risk of lung cancer mortality based on ALARM-NS for an average risk or high risk never smoker, for combinations of age and lung function (FEV1/FVC). An average risk profile is defined as having the average covariate value for all predictors other than those varied, and a high risk profile is defined as having the highest risk covariate value observed in the CKB (based on direction of effect) for all predictors other than those varied.

We compared our absolute risk models to the recently-updated USPSTF criteria for lung cancer screening (see **Table 3**). Using the USPSTF criteria, 35.5% of CKB ever-smokers would be eligible for lung cancer screening. To compare our model to the USPSTF criteria, we applied the five-year risk threshold that produced an equivalent specificity to the USPSTF criteria - our model (ALARM-ES) demonstrated an improved sensitivity (82.1% vs. 68.6% based on USPSTF), positive predictive value (1.1% vs. 1.0%), and negative predictive value (97.8% vs. 96.3%), while selecting a similar proportion of the population for screening. At a five-year risk threshold that has an equivalent sensitivity to the USPSTF criteria, ALARM-ES would select 11.5% fewer ever-smokers for screening (24.0% vs. 35.5% based on USPSTF).

**Table 3.** Comparison of ALARM-ES and LCDRAT against the United States Preventive Services Task Force (USPSTF) 2021 criteria for lung cancer screening. The USPSTF recommends annual screening for adults age 50-80 years, with 20 pack-year smoking history, and smoking cessation less than 15 years prior for former smokers. We compared the ever-smoker models two ways: (1) using a risk threshold that matches the specificity of USPSTF criteria, and (2) using a risk threshold that matches the sensitivity of USPSTF criteria.

|  | **Specificity** | **Sensitivity** | **PPV** | **NPV** | **% eligible for screening** |
| --- | --- | --- | --- | --- | --- |
| USPSTF-2021 | 65.5% | 68.6% | 1.0% | 96.3% | 35.5% |
| **ALARM-ES** |  |  |  |  |  |
| Matched on specificity | Same as USPSTF | 82.1% | 1.1% | 97.8% | 36.4% |
| Matched on sensitivity | 77.8% | Same as USPSTF | 1.5% | 97.1% | 24.0% |
| **LCDRAT** |  |  |  |  |  |
| Matched on specificity | Same as USPSTF | 82.2% | 1.1% | 97.7% | 36.5% |
| Matched on sensitivity | 78.0% | Same as USPSTF | 1.4% | 97.0% | 23.9% |

Abbreviations: ALARM = Asian Lung cancer Absolute Risk Model; LCDRAT = Lung Cancer Death Risk Assessment Tool; NPV = Negative predictive value; PPV = Positive predictive value; USPSTF = United States Preventive Services Task Force, USPSTF.
Note: Time-dependent sensitivity, specificity, PPV, and NPV for USPSTF-2021 and ALARM-ES were based on inverse-probability of censoring weighted (IPCW) estimates for a cumulative/dynamic definition of cases and control proposed by Blanche *et al.* (2013). For LCDRAT, the IPCW estimates were based on the time-dependent Sens/Spec/PPV/NPV proposed by Blanche *et al.* (2013) which extends the cumulative/dynamic definition of cases and control for the competing risks setting.

We applied the established Lung Cancer Death Risk Assessment Tool (LCDRAT) (20) to the CKB ever-smokers to assess how our model (ALARM-ES) performed when compared against a validated ever-smoker lung cancer mortality model. Details of how LCDRAT was applied and evaluated in the CKB are described in the Supplemental Methods. Calibration and discrimination statistics are presented in **Supplemental Table 3**. LCDRAT and LCDRAT-Constrained had similar discriminative performance (i.e., AUC) as ALARM-ES, however, ALARM-ES had superior calibration. The expected/observed death ratios were 1.04 (95% CI: 0.92-1.20) in ALARM-ES and 1.57 (95% CI: 1.46-1.69) in LCDRAT, and the differences between expected and observes deaths (per 100,000) were 18.68 (−45.68-89.53) and 288.02 (95% CI: 250.10-326.47), respectively. This is not surprising as risk models developed in North American cohorts may not be well calibrated in Asian populations, which are reflected in the calibration statistics, despite good overall discrimination.

## Discussion

We established risk prediction models based on a relatively simple set of predictors that can be ascertained during a routine physician visit, which can accurately discriminate high and low risk individuals for lung cancer mortality in an Asian population better than the US-based current recommended screening guidelines. These are some of the first prediction models for lung cancer mortality that were developed and evaluated specifically for an Asian population and separately for never and ever-smokers. Our model has shown superior calibration compared to the established lung cancer mortality model (LCDRAT), while maintaining a comparable model accuracy. At a risk threshold that matches the specificity of the USPSTF criteria, ALARM-ES has better sensitivity, PPV, and NPV than USPSTF criteria, while selecting a similar proportion of the population for lung cancer screening.

Despite the global reductions in smoking prevalence in most parts of the world (21), lung cancers among never-smokers is an increasingly important public health concern (22). The proportion of lung cancers in never smokers has been increasing and lung cancer in never smokers is one of the most common cancers (23). It is estimated that the epicenter of lung cancer in the next few decades would be within Asian countries, where a substantial proportion of lung cancers would occur in never smokers. This highlights the importance of having risk models developed specifically for this population. The International Association of Lung Cancer Study (IASLC) recently released a statement specifically focused on never smokers, in which it emphasizes the importance of risk-based screening for never smokers (22). While widespread screening of never-smokers at the current stage would not be effective, the development and validation of risk predictions will play a critical role (22). The report encourages and recommends modelling studies focused on lung cancer risk estimation for never-smokers in order to eventually realize the benefit of risk-based screening in this group (22).

Furthermore, it is well known that lung cancers occurring in never smokers represent a distinct form of the disease (23,24). Lung cancers occurring among never-smoker are more commonly adenocarcinomas, with higher proportions of EGFR mutations, which makes them highly targetable by therapeutics leading to improved prognosis (25); although these therapeutic agents are typically administered when the disease is at a later, incurable stage, the majority of lung cancer patients with EGFR-mutations will eventually become incurable at some point in their cancer course. This contributes, in part, to the improved survival observed among never-smokers (25). Based on recent findings from the ADAURA trial, there should be an even stronger push to identify these cancers at an earlier resectable stage, where the future use of EGFR tyrosine kinase inhibitors can improve disease free survival (26). In the absence of primary smoking history as a risk factor, statistical models based on a constellation of risk factors will be required in order to identify high-risk never-smokers for screening. The ability to identify and screen high-risk never-smokers for lung cancer is an important public health concern.

In this study, we developed and evaluated risk models for estimating the five-year absolute risk of lung cancer mortality in the hold-out test set. As lung cancer incidence data was not yet available from CKB at the time of our analysis, lung cancer mortality was modeled as the endpoint. However, given that mortality is the primary endpoint of CT screening programs, modeling lung cancer mortality as the outcome also has the clinical advantage of identifying individuals who are at high risk of lung cancer death, and in return, reduces the possibility of over-diagnosis.

Note that in the competing risk setting, the absolute risk depends on both cause-specific hazards and cause-specific hazard ratios only reflect how a variable affects the rate at which the cause-specific event occurs and might not directly coincide with how that variable affects the probability of that event occurring, since the absolute risk depends on the cause-specific hazards for lung cancer mortality and all-other cause mortality. Still, we may draw some conclusions about the cause-specific hazard models fitted in this study.

Lung cancers have previously been found to affect never-smoking women disproportionately more than men (25,27), though women are still observed to have a survival advantage (28), which is consistent with our findings. Due to limitations in the data, we were unable to determine a participants family history of lung cancer, so we used family history of any cancer, which is expected to be a relatively weak proxy. Exposure to cooking fuel has previously been identified as a risk factor for lung cancer (29,30), particularly among Asian women (31,32). However, in our study we did not observe an association between cooking fuel exposure and lung cancer mortality. We believe this may be due, in part, to imprecise measurements of this putative risk factor. Similar to passive tobacco exposure (i.e., second-hand smoke), cooking fuel exposure is subject to recall bias and thus it is difficult to accurately measure cumulative lifetime exposure.

We identified four previous studies which developed or adapted risk models for never-smokers, and two were based in Asian populations. The PLCOall2014 model was developed in both ever and never-smoker population and was adapted for use in never-smokers by removing smoking-related predictors (33). We applied the PLCOall2014 to the CKB, and our model achieved higher AUC for never smokers (0.77 versus 0.76 for PLCOall2014) and ever-smokers (0.81 vs. 0.75 based on PLCOall2014). Details of this comparison are described in the Supplementary Methods. A second study was developed in a predominantly Caucasian cohort (UK Biobank) (34). A third study was developed in a Taiwanese cohort and models were built separately for never, light, and heavy smokers (35). This study achieved a training-set AUC of 0.806 (95% CI: 0.790-0.819) among never-smokers but included several serological protein biomarkers unavailable in the CKB that are not routinely tested in health populations and therefore could not be validated. The authors presented limited evidence of model calibration across risk groups and by smoking status. A fourth model was developed for never-smoker females only in an Asian case-control study and achieved an AUC of 0.71 (95% CI: 0.66-0.77) (36). However, this study was not based on prospective data, and would require the collection of genetic data (i.e., genotyping) which may be prohibitive for widespread application. The model without the 9 genetic variants performed moderately worse (AUC=0.69, 95% CI: 0.64-0.75).

Based on ALARM-NS, fewer than 1% of the never-smokers in CKB reached a five-year risk threshold of at least 1.5% to be eligible lung cancer screening, with the highest-risk never smoker achieving a risk of 2.7%. The marginal age-specific lung cancer and other cause mortality rates for men and women were lower in the CKB than those observed in the general Chinese population, based on Global Burden of Disease 2019 mortality estimates (see Supplemental Figure 4). This may be due, in part, to a “healthy volunteer” effect. This phenomenon occurs when those persons who volunteer for a study are healthier and not fully representative of the general population. As such, we anticipate the actual risk distribution in the general Chinese population would be higher than what is observed in CKB (Supplementary Figure 1). It is possible that potential measurement error of important predictors (e.g., indoor air pollution) has led to an underestimate of absolute risks. A larger proportion of individuals would be at a higher risk for lung cancer mortality and may be eligible for screening based on applying our model to the general Chinese population.

In summary, we developed absolute risk models that accurately identify both ever- and never-smokers at high-risk for lung cancer-related mortality, accounting for the competing risk of all-other-cause mortality. These models were developed exclusively using prospective data collected in a large Asian population, and models were fitted separately based on smoking history (ALARM-NS and ALARM-ES). Our models discriminated high and low risk for lung cancer death similarly well to an established lung cancer mortality model (i.e., LCDRAT), but demonstrated much better absolute risk calibration for an Asian population. We made our models freely accessible as a user-friendly R package (https://github.com/mattwarkentin/ALARM). In the future, these models may be useful for identifying a subset of the Asian population at high-risk for lung cancer mortality who may benefit from lung cancer screening. The next step will be to externally validate our models in an independent cohort.

## Data Availability

The data is available from the China Kadoorie Biobank (https://www.ckbiobank.org/) upon approval of the CKB Data Access Committee.

## Notes

### Data Availability Statement

All of the data used in the completion of this study are publicly available, by request, from the China Kadoorie Biobank Data Access Committee (https://www.ckbiobank.org/site/Data+Access).

The absolute risk models presented in this article are made available as a user-friendly, free and open-source tool, at the following link: https://github.com/mattwarkentin/ALARM.

### Contributors

RJH conceived the study and obtained the funding. MTW and RJH contributed equally to the design of the study and co-led the writing of the article. MTW led the statistical analysis with scientific input from RJH, OEG, and MCT. All coauthors contributed to the result interpretations, critical review of the manuscript, and approval of the final version.

