## Supplementary Materials for "Asian Lung Cancer Absolute Risk Models for lung cancer mortality based on China Kadoorie Biobank"

#### Contents

**Supplementary Methods** **2**

Asian Lung Cancer Absolute Risk Model (ALARM) to predict *t*-year risk of lung cancer mortality 2

Applying the LCDRAT model to the China Kadoorie Biobank 4

Applying the PLCOall2014 model to the China Kadoorie Biobank 6

**References** **8**

### Supplementary Methods

#### Asian Lung Cancer Absolute Risk Model (ALARM) to predict *t*-year risk of lung cancer mortality

Our absolute risk models for lung cancer mortality account for competing forces of mortality and are a function of both the hazard of mortality due to lung cancer and the hazard of mortality due to all other causes. Thus, we fit three separate cause-specific hazard models to form the two absolute risk models for never and ever smokers (i.e. ALARM). Each cause-specific hazard model was estimated based on flexible parametric survival models. The never-smoker absolute risk model is based on a never smoker-specific model for lung cancer mortality and an all-other cause mortality model. The ever-smoker absolute risk model is based on an ever smoker-specific model for lung cancer mortality and an all-other cause mortality model. The same other cause mortality model was used for both absolute risk models.

In the competing risk setting, the risk of death from lung cancer at time *t* (i.e. the cumulative incidence function) is a function of both the cause-specific hazard for lung cancer mortality and the cause-specific hazard for other cause mortality (1,2). In the equation below, $\lambda_1(t|x_1)$ is the cause-specific hazard for lung cancer mortality conditional on covariates $x_1$, and $\lambda_2(t|x_2)$ is the cause-specific hazards for other cause mortality conditional on covariates $x_2$. Both $\lambda_1$ and $\lambda_2$ were estimated based on flexible parametric survival models. $\lambda_1$ were fitted separately and are different for never and ever smokers, while the same other cause mortality model is used for $\lambda_2$.

In a general form, the absolute risk model (i.e., cumulative incidence function) is defined as:

$$\text{CIF}(t|x_1, x_2) = \int_0^t \lambda_1(u|x_1)\,\text{exp}\left[-\int_0^u \lambda_1(v|x_1) + \lambda_2(v|x_2)dv\right]du \quad (1)$$

For never-smokers, **ALARM-NS** is defined as:

$$\text{ALARM-NS}(t|x_1, x_2) = \int_0^t \lambda_1^{\text{NS}}(u|x_1)\,\text{exp}\left[-\int_0^u \lambda_1^{\text{NS}}(v|x_1) + \lambda_2(v|x_2)dv\right]du \quad (2)$$

For ever-smokers, **ALARM-ES** is defined as:

$$\text{ALARM-ES}(t|x_1, x_2) = \int_0^t \lambda_1^{\text{ES}}(u|x_1)\,\text{exp}\left[-\int_0^u \lambda_1^{\text{ES}}(v|x_1) + \lambda_2(v|x_2)dv\right]du \quad (3)$$

#### Applying the LCDRAT model to the China Kadoorie Biobank

We applied the Lung Cancer Death Risk Assessment Tool (LCDRAT) to the China Kadoorie Biobank to assess how well it performs in a new population. LCDRAT is a lung cancer mortality model for ever-smokers, developed in the control arm of the Prostate, Lung, Colorectal, and Ovarian Cancer Screening Trial (PLCO) (3). LCDRAT is based on cause-specific Cox models fitted separately for lung cancer death and deaths from all other causes (i.e. competing risks). The five-year probability of dying of lung cancer (in the absence of CT screening) is estimated as a function of both cause-specific hazards models to account for the competing risks of death using the following equation (adapted from Katki *et al*. (3)):

$$R_1 = \int_0^5 \lambda_1(u; x_1) \exp\left(-\int_0^u (\lambda_1(v; x_1) + \lambda_2(v; x_2))dv\right) du \qquad (4)$$

Where $\lambda_1(t)$ is the cause-specific hazard for lung cancer death, $\lambda_2(t)$ is the cause-specific hazard for death from all other causes, and $x_1$ and $x_2$ are the covariates for each of the cause-specific Cox models, respectively. Both models included the following covariates: age, education, sex, race, smoking intensity, smoking duration, years since quitting smoking, body mass index, and self-reported history of emphysema. The lung cancer death model further included family history of lung cancer in first-degree relatives. A simplified (i.e. fewer predictors) version of the LCDRAT (hereafter referred to as LCDRAT Constrained) was also evaluated. LCDRAT Constrained excludes race, education, BMI, history of emphysema, and family history of lung cancer.

We used the `R` package `lcmodels` (available at: https://dceg.cancer.gov/tools/risk-assessment/lcmodels) to estimate the five-year probability of dying of lung cancer in the absence of screening using both LCDRAT and LCDRAT Constrained. We evaluated the performance of these models by the time-dependent area under the ROC curve (i.e. discrimination) and by comparing the expected and observed number of deaths (i.e. calibration). The time-dependent AUC was estimated using the `timeROC` package based on the definition of the Cumulative/Dynamic time-dependent ROC in the competing risks setting as proposed by Blanche *et al* (5). Expected (Exp) number of deaths (per 100,00) were calculated based on the mean model-predicted risks multiplied by 100,000, and the observed (Obs) number of deaths were based on the value of the non-parametric Aalen-Johansen estimator for competing risks at five-years multiplied by 100,000. The ratio of expected to observed ($\mathtt{Exp}/\mathtt{Obs}$) and the difference between expected and observed ($\mathtt{Exp} - \mathtt{Obs}$) were also calculated. Percentile-based 95% confidence intervals were constructed using 500 bootstrap resamples.

#### Applying the PLCOall2014 model to the China Kadoorie Biobank

We evaluated how well the established PLCOall2014 model performed in the China Kadoorie Biobank cohort. The PLCOall2014 model was developed in a North American cohort of never and ever smokers (6). PLCOall2014 is an adapted version of the PLCOm2012 model (7) to allow for the inclusion of never smokers. This model includes the following predictors: age, education, body mass index (BMI), history of COPD, race/ethnicity (White, Black, Hispanic, Asian, Native Hawaiian or Pacific Islander, American Indian or Alaskan Native), personal history of cancer (PHC), family history of lung cancer (FHLC), and the smoking variables (which do not apply to never-smokers) smoking status (former or current), average number of cigarettes per day (intensity), years smoked (duration), and years since quitting (quit time). The model was fitted as a logistic regression for estimating 6-year risk of lung cancer incidence. The equation for the PLCOall2014 model is shown below.

Our model is a lung cancer death model, and so we cannot directly compare the predictions made from the two models. Additionally, since we do not have data on lung cancer incidence, we cannot assess the calibration of the PLCOall2014 model in the CKB. Thus, we compared the area under the receiver operating characteristic curve (AUC) achieved by the PLCOall2014 model to the time-dependent AUCs achieved by our models. Since the PLCOall2014 model includes FHLC, which is not reported in the CKB, we could only apply the model to those in CKB with no family history of any cancer (83% of the total CKB), who, by definition, must also not have a FHLC. For computing the AUC, we constructed the binary response variable by considering all participants who died from lung cancer before 6 years as cases and all other participants considered as controls.

$$
\begin{aligned}
Logit_{PLCOall2014} = & -7.02198 + (0.079597 * (\texttt{Age} - 62)) - \\
& (0.0879289 * (\texttt{Education} - 4)) - (0.028948 * (\texttt{BMI} - 27)) + \\
& (0.3457265 * \texttt{COPD}) + (0.3211605 * \texttt{Black}) - \\
& (0.8203332 * \texttt{Hispanic}) - (0.5241286 * \texttt{Asian}) - \\
& (0.952699 * \texttt{Pacific Islander}) + (1.364379 * \texttt{Native American}) + \\
& (0.4845208 * \texttt{PHC}) + (0.5856777 * \texttt{FHLC}) + \\
& (2.542472 * \texttt{Former}) + (2.799727 * \texttt{Current}) + \\
& (0.0305566 * (\texttt{Duration} - 27)) - (0.0321362 * (\texttt{Quit Time} - 10)) - \\
& (0.1815486 * (((\texttt{Intensity}/100)^{-}1) - 4.021541613))
\end{aligned}
\tag{5}
$$

$$
P(\texttt{6 year risk of lung cancer}) = \frac{e^{Logit_{PLCOall2014}}}{1 + e^{Logit_{PLCOall2014}}} \tag{6}
$$

**Supplemental Table 1.** Distribution of demographic characteristics of China Kadoorie Biobank based on smoking status. Means and standard deviations are reported for numeric variables, and frequencies and proportions for categorical variables.

| | Never Smokers No. (%) | Ever Smokers No. (%) |
|---|---|---|
| N | 346,617 | 166,022 |
| Age (years) | | |
| mean [sd] | 51.4 [10.6] | 53.2 [10.7] |
| Length of follow-up (years) | | |
| mean [sd] | 10.0 (1.7) | 9.7 [2.1] |
| Sex | | |
| Female | 292,700 (84%) | 9,797 (6%) |
| Male | 53,917 (16%) | 156,225 (94%) |
| Family history of cancer | | |
| No | 288,612 (83%) | 136,720 (82%) |
| Yes | 58,005 (17%) | 29,302 (18%) |
| Personal cancer history | | |
| No | 344,849 (99%) | 165,287 (>99%) |
| Yes | 1,768 (1%) | 735 (<1%) |
| Emphysema or bronchitis | | |
| No | 319,519 (92%) | 147,246 (89%) |
| Yes | 27,098 (8%) | 18,776 (11%) |
| Smoking intensity (cigs/day) [a] | | |
| mean [sd] | — | 17.8 [10.8] |
| Smoking duration (years) [a] | | |
| mean [sd] | — | 28.5 [11.6] |
| Years since cessation (years) [b] | | |
| mean [sd] | — | 8.8 [8.5] |
| FEV1/FVC (%) | | |
| mean [sd] | 85.0 [8.1] | 83.4 [9.4] |
| Body mass index (kg/m$^2$) | | |
| mean [sd] | 23.8 [3.4] | 23.3 [3.3] |
| Household income [c] | | |
| <2500 yuan | 9,843 (3%) | 5,694 (3%) |
| 2,500-4,999 yuan | 22,944 (7%) | 11,680 (7%) |
| 5,000-9,999 yuan | 65,299 (19%) | 29,262 (18%) |
| 10,000-19,999 yuan | 102,022 (29%) | 46,910 (28%) |
| 20,000-34,999 yuan | 85,894 (25%) | 40,787 (25%) |
| ≥35,000 yuan | 60,615 (17%) | 31,689 (19%) |
| Cooking fuel exposure [d] | | |
| None | 99,693 (29%) | 104,683 (63%) |
| Low | 47,489 (14%) | 33,339 (20%) |
| Medium | 131,132 (38%) | 19,119 (12%) |
| High | 68,303 (20%) | 8,881 (5%) |

Abbreviations: FEV1 = forced-expiratory volume, 1-second; FVC = forced vital capacity; sd = standard deviation.

[a] Average smoking intensity and number of years smoking (duration) includes current and former smokers; never smokers are excluded.

[b] Years since smoking cessation only includes former smokers; current and never smokes are excluded.

[c] Approximate equivalents in USD ($), rounded to the nearest dollar: <384, 384-767, 767-1,537, 1,537-3,075, 3,075-5,381, and ≥5381.

[d] Cooking fuel exposure groups were formed based on 25$^{th}$ and 75$^{th}$ percentiles of the cumulative cooking exposure distribution. Cumulative cooking exposure was calculated based on the self-reported frequency of exposure to potentially harmful cooking fuels (i.e., coal, wood, or other, as compared to gas or electricity).

**Supplemental Table 2.** Estimates from all-other cause mortality flexible parametric survival model. All-other cause-specific hazard ratios with 95% confidence intervals and beta coefficients (log hazard ratios and standard errors) are reported.

| | **HR (95% CI)** | **Beta (SE)** |
|---|---|---|
| Age (years) | 1.10 (1.10-1.10) | 0.0975 (0.0007) |
| Sex (Female vs. Male) | 0.71 (0.68-0.73) | -0.3459 (0.0188) |
| Smoking status | | |
| Never smoker | — | — |
| Former smoker | 1.43 (1.36-1.49) | 0.3543 (0.0231) |
| Current smoker | 1.41 (1.36-1.47) | 0.3461 (0.0197) |

Abbreviations: CI, confidence interval; HR, hazard ratio; SE, standard error.

**Supplemental Table 3.** Discrimination and calibration statistics for ALARM-ES, ALARM-NS, LCDRAT, and LCDRAT Constrained in the China Kadoorie Biobank (CKB). Performance metrics are based on the hold-out test set for ALARM-ES and ALARM-NS, while LCDRAT and LCDRAT-Constrained were applied to the entire eligible set of ever-smokers in the CKB.

| | **Number of expected deaths (per 100,00)** | **Number of observed deaths (per 100,000)** | **Expected / Observed (95% CI)** | **Expected - Observed (95% CI)** | **Area Under the Curve (95% CI)** |
|---|---|---|---|---|---|
| **ALARM-ES** [a] | 536.83 (530.46-542.91) | 518.15 (448.25-583.25) | 1.04 (0.92-1.20) | 18.68 (-45.68-89.53) | 0.81 (0.79-0.84) |
| **ALARM-NS** [a] | 157.97 (156.82-159.04) | 154.69 (131.60-180.09) | 1.02 (0.88-1.20) | 3.28 (-22.24-26.58) | 0.77 (0.73-0.81) |
| **LCDRAT** [b] | 793.62 (786.88-800.09) | 505.61 (468.64-542.20) | 1.57 (1.46-1.70) | 288.02 (250.10-326.47) | 0.81 (0.80-0.82) |
| **LCDRAT-Constrained** [b,c] | 835.12 (828.36-841.70) | 505.61 (468.95-541.47) | 1.65 (1.53-1.78) | 329.52 (294.38-366.17) | 0.81 (0.79-0.82) |

Abbreviations: ALARM = Asian Lung Cancer Absolute Risk Model; CI = confidence interval; ES = ever smoker; LCDRAT = Lung Cancer Death Risk Assessment Tool; NS = never smoker.

Note: Bootstrap-based 95% confidence intervals were constructed using the percentile-based approach (i.e. by taking the 2.5$^{th}$ and 97.5$^{th}$ percentile of the bootstrap distribution for a given statistic).

[a] Expected deaths (per 100,000) are based on the average model-predicted risk. Observed deaths (per 100,000) are based on the complement of the Kaplan-Meier survival function. Time-dependent area under the curve is based on the inverse probability of censoring weighted (IPCW) estimator for right-censored survival data using the cumulative/dynamic definition of cases and control proposed by Blanche *et al.* (2013).

[b] LCDRAT and LCDRAT-Constrained are only applied to ever-smokers in the CKB. In addition, due to the absence of data collected on family history of lung cancer, we only apply the model to participants with no family history of any cancer, who, by definition, must not have a family history of lung cancer. Expected deaths (per 100,000) are based on the average model-predicted risk. Observed deaths (per 100,000) are based on the non-parametric Aalen-Johansen estimator for competing risks. Time-dependent area under the curve is based on the inverse probability of censoring weighted (IPCW) estimator for right-censored competing risks data using the cumulative/dynamic definition of cases and control proposed by Blanche *et al.* (2013).

[c] LCDRAT Constrained excludes race, education, body mass index, history of emphysema, and family history of lung cancer when compared to LCDRAT.

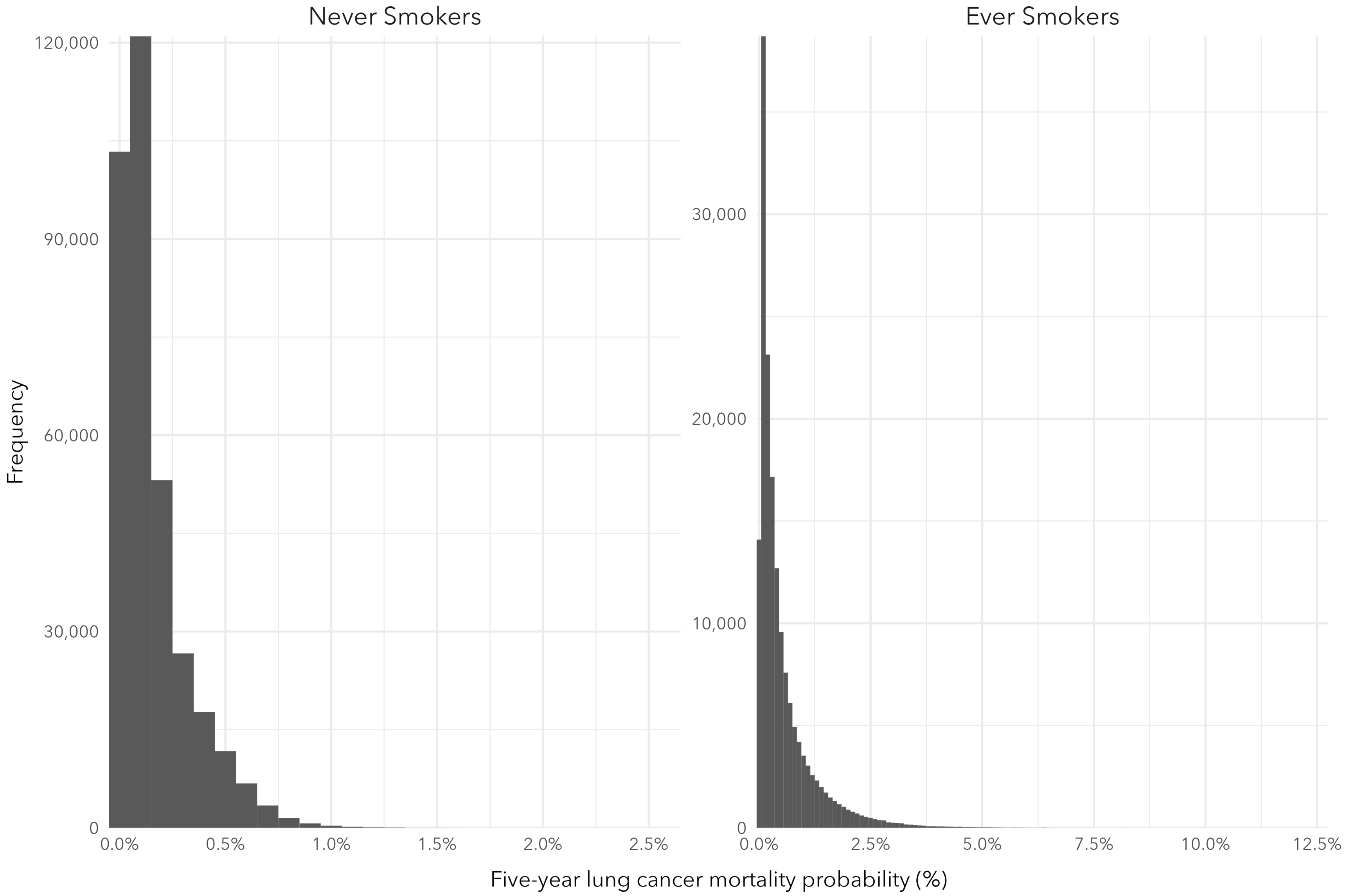


**Supplemental Figure 1.** Distributions of model-predicted five-year absolute risks of lung cancer mortality for ALARM-NS (never-smokers) and ALARM-ES (ever-smokers) among all participants in the China Kadoorie Biobank.

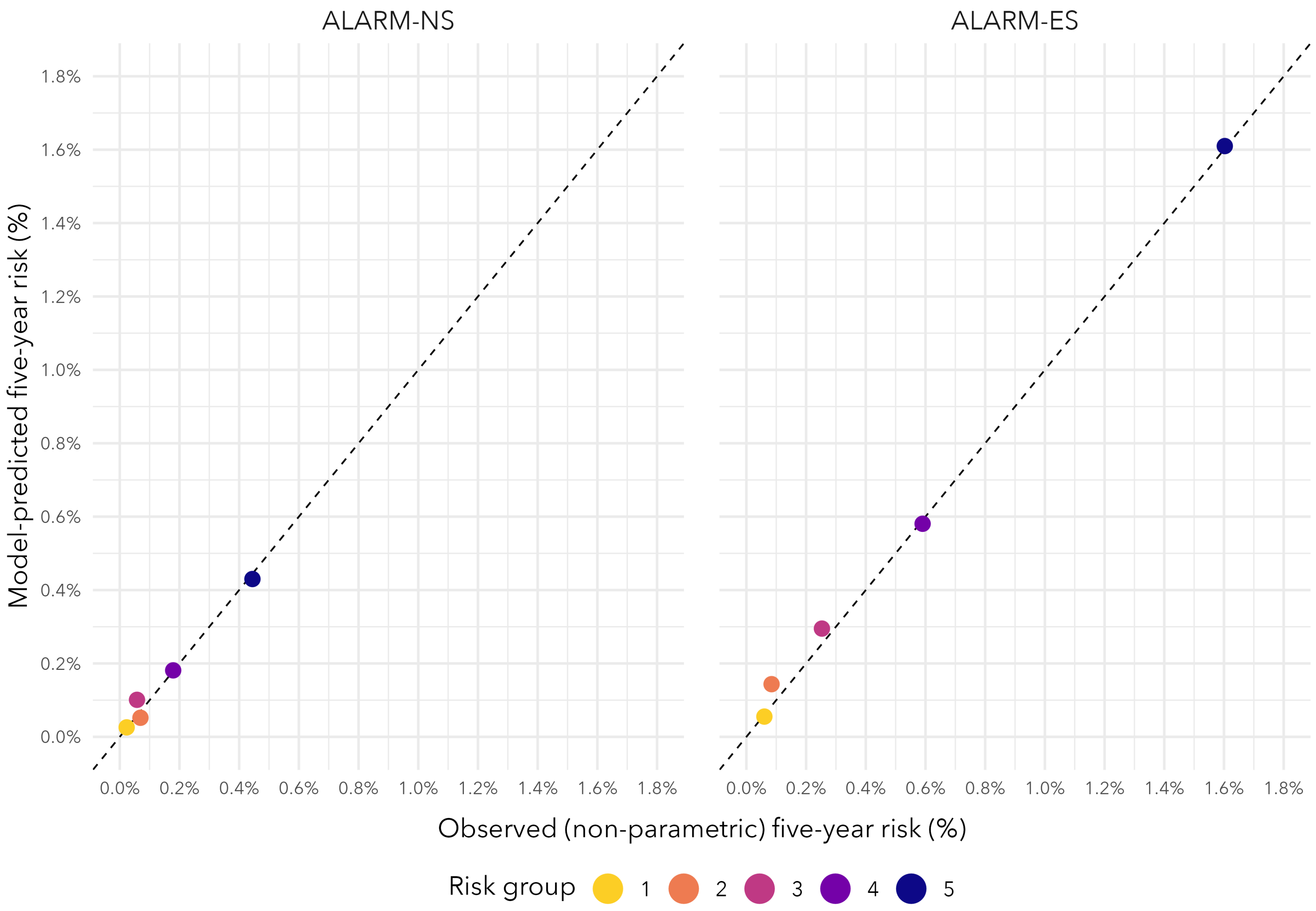


**Supplemental Figure 2.** Calibration plot comparing the average model-predicted five-year absolute risks to the observed five-year risks (based on the non-parametric Aalen-Johansen estimator) across risk groups, separately for ALARM-NS (never-smokers) and ALARM-ES (ever-smokers).

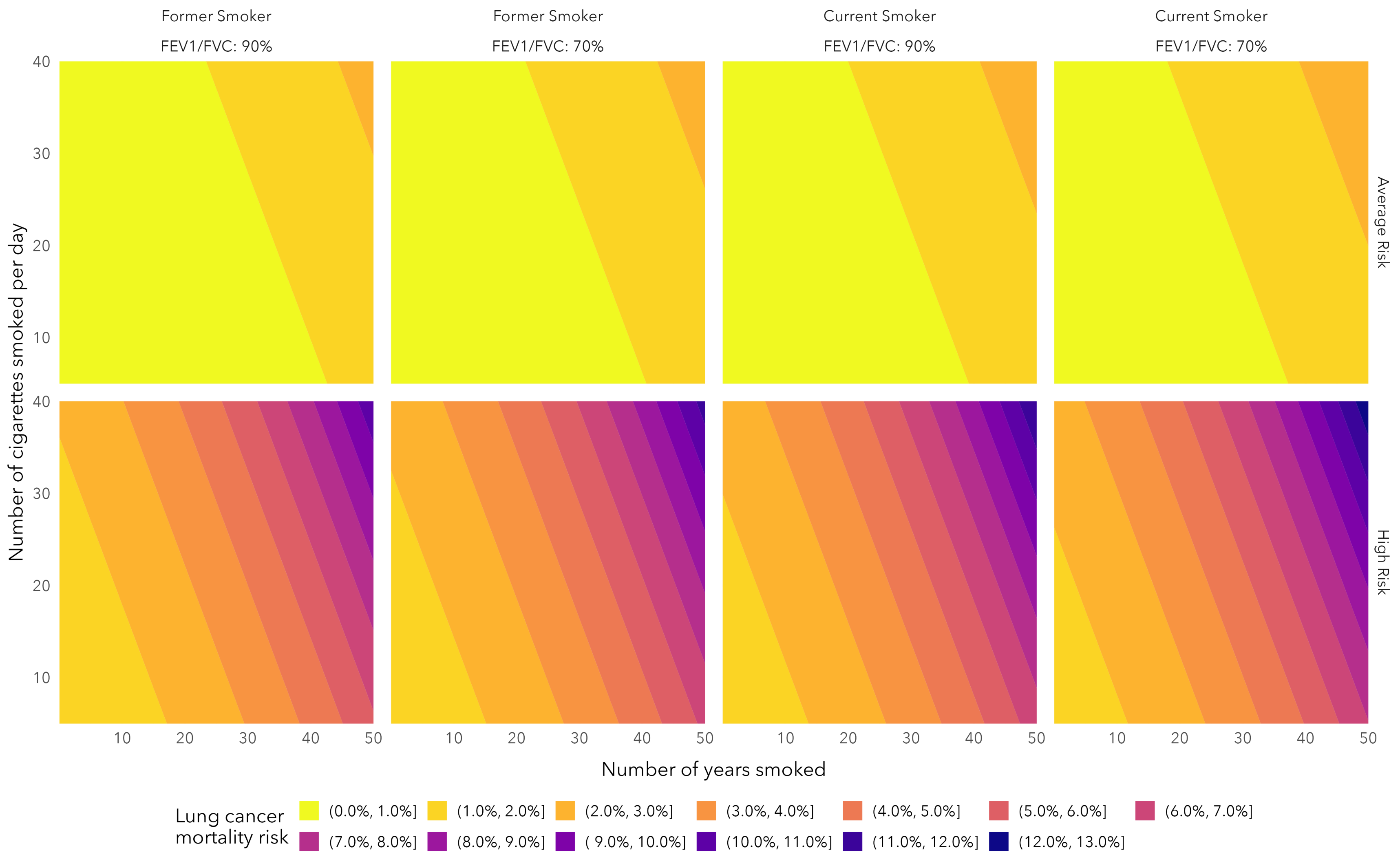


**Supplemental Figure 3.** Five-year absolute risk of lung cancer mortality based on ALARM-ES for a 70 year old average risk or high risk current or former smoker for combinations of smoking intensity, smoking duration, and FEV1/FVC. An average risk profile is defined as having the average covariate value for all predictors other than those varied, and a high risk profile is defined as having the highest risk covariate value observed in the CKB (based on direction of effect) for all predictors other than those varied.

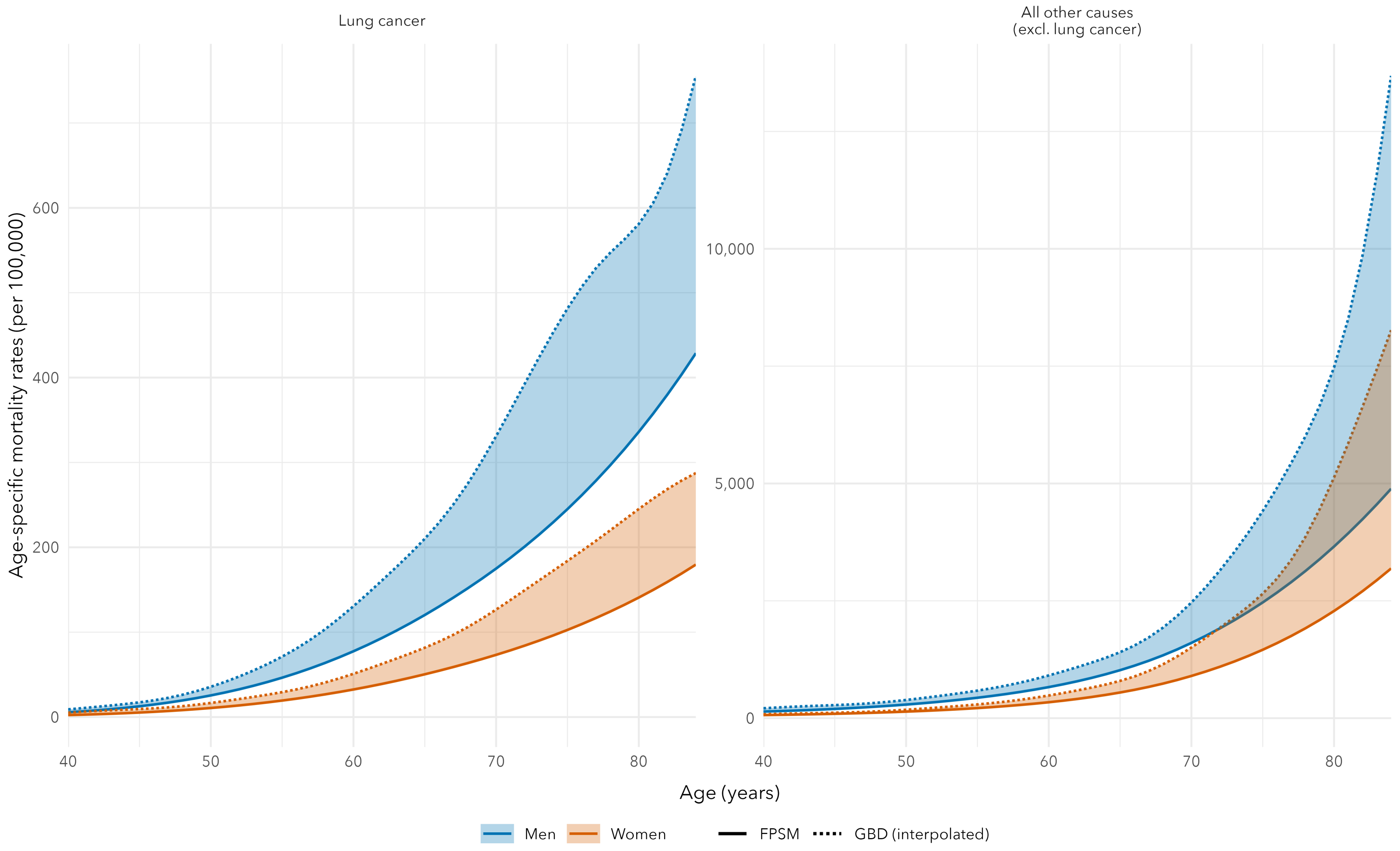


**Supplemental Figure 4.** Age-specific lung cancer and other cause mortality rates for men and women (per 100,000). Solid lines are based on flexible parametric survival models (FPSM) estimated using China Kadoorie Biobank, while the dashed lines are based on Global Burden of Disease (GBD) 2019 estimates for China. GBD estimates were reported in five-year age bins. Mortality rates for integer ages were interpolated using natural cubic splines passing through the mid-point of age bins. The filled area between lines indicate the differences in age-specific rates between CKB and the general Chinese population.
